# Criteria to Achieve Safe Antimicrobial Intravenous-to-Oral Switch in Hospitalised Adult Populations: A Systematic Rapid Review

**DOI:** 10.1101/2022.09.01.22279505

**Authors:** Eleanor J Harvey, Monsey McLeod, Caroline De Brún, Diane Ashiru-Oredope

**Affiliations:** Healthcare-Associated Infection (HCAI), Fungal, Antimicrobial Resistance (AMR), Antimicrobial Use (AMU) & Sepsis Division, United Kingdom Health Security Agency (UKHSA), London, UK; Antimicrobial Prescribing and Medicines Optimisation, NHS England & NHS Improvement, London, UK; Knowledge and Library Services, United Kingdom Health Security Agency (UKHSA), London, UK; School of Pharmacy, University of Nottingham, Nottingham, UK

**Author notes:** Corresponding Author Diane Ashiru-Oredope.

## Abstract

**Introduction:** Antimicrobial stewardship and patient safety strategies include early intravenous-to-oral switch (IVOS) for antimicrobials.

**Aim:** This rapid review aimed to assess and collate IVOS criteria from the literature to achieve safe and effective antimicrobial IVOS in the hospital inpatient adult population.

**Method:** The rapid review follows the PRISMA statement and is registered with PROSPERO. Systematic literature searches were conducted. Articles of adult populations published between 2017-2021 were included. IVOS criteria from UK hospital IVOS policies were categorised to inform the framework synthesis of the literature criteria.

**Results:** IVOS criteria from 45/164 (27%) UK IVOS policies were categorised into a 5-section framework: 1-Timing of IV antimicrobial review, 2-Clinical signs and symptoms, 3-Infection markers, 4-Enteral route, and 5-Infection exclusions. The literature search identified 477 papers, of which 16 were included. The most common timing for review was 48-72 hours from initiation of intravenous antimicrobial (n=5, 30%). Nine studies (56%) stated clinical signs and symptoms must be improving. Temperature was the most frequently mentioned infection marker (n=14, 88%). Endocarditis had the highest mention as an infection exclusion (n=12, 75%). Overall, 33 IVOS criteria were identified to go forward into the Delphi process.

**Conclusion:** Through the rapid review, 33 IVOS criteria were collated and presented within 5 distinct and comprehensive sections. The literature highlighted the possibility of reviewing IVOS before 48-72 hours, and of presenting HR, BP and RR as a combination early warning score criterion. The criteria identified can serve as a starting point of IVOS criteria review for any institution globally, as no country or region limits were applied. Further research is required to achieve consensus on IVOS criteria from healthcare professionals that manage patients with infections.

**What is already known on this topic:** Antimicrobial intravenous-to-oral switch has benefits such as decreased risk of catheter-related infections, reduced equipment costs and increased patient mobility and comfort. Acute hospitals often develop and implement individualised IVOS policies with varying levels of evidence base.

**What this study adds:** This study provides evidence-based IVOS criteria to standardise practice in hospital settings.

**How this study might affect research, practice or policy:** IVOS criteria can be taken forward through a consensus process with healthcare professionals providing the care for hospitalised adult patients and making the decisions regarding infection management. In the acute hospital setting, IVOS criteria can be operationalised to promote best practice. The criteria can also be considered as part of organisation-wide audits and quality/policy incentives.

## 1 Introduction

Antimicrobial resistance (AMR) is a public health threat,^1^ and tackling this threat requires joint efforts at local, national and global levels across all care settings.^2^ In the acute hospital care setting, antibiotic consumption is known to promote resistant and multidrug-resistant bacteria leading to healthcare-associated infections.^3^ Antibiotic prescribing in England continued to rise in acute hospitals between 2016 and 2020, with specialist hospitals accounting for the highest increase of 31.2%.^4^

Antimicrobial stewardship (AMS) is defined as a set of strategies to promote the responsible use of antimicrobials^5^ for the purpose of protecting public health.^6^ Across secondary care, the COVID-19 pandemic had a negative impact on AMS activities, but evidence suggests the pandemic also encouraged collaboration between infection specialists and non-specialists to strengthen activities going forward.^7^ Antimicrobial intravenous-to-oral switch (IVOS) has been classified as a “low-hanging fruit” AMS activity; in other words, an attainable AMS strategy despite constrained limitations.^8^ The literature outlines numerous benefits for IVOS; including decreased risk of catheter-related infections, reduced equipment costs and increased patient mobility and comfort.^9, 10^ Additionally, decreased medical equipment usage has the potential to reduce hospital’s carbon footprint.^11^ In clinically stable patients, studies report that a timely IVOS is safe and of equal efficacy to the full course of IV therapy,^12^ with no negative reports on patient outcome.^12^ In the United Kingdom (UK), the national Start Smart – Then Focus AMS Toolkit recommends IVOS as an important outcome following review of prescribed antimicrobial within 48-72 hours.

Acute hospitals have developed and implemented their own individualised local IVOS policies and there is currently no standardised national criteria recommended for IVOS in the UK. The evidence-base behind the criteria included in the individualised policies is largely unknown. Additionally, they include a variety of criteria themes and nuances in criterion wording. The standardisation of clinical guidance, whether protocols, checklists or other, has led to enhanced patient safety and outcomes and driven improved quality of care.^13-15^

This rapid review aims to evaluate the evidence-base for IVOS criteria and collate criteria into common themes. This review will subsequently inform a Delphi consensus-gathering process, led by the UK Health Security Agency (UKHSA), to establish standardised, evidence-based, national antimicrobial IVOS criteria. The UK has previously never had nation-wide IVOS criteria available for use by hospitals, this review will form the basis for included IVOS criteria.

## 2 Method

### 2.1 Protocol and registration

This review was informed by the World Health Organisation^16^ and Cochrane^17^ rapid review protocols, and follows the Preferred Reporting Items for Systematic Reviews and Meta-Analyses (PRISMA) 2020 item checklist (**Supplementary Table S1**) and guidance.^18,19^ The review protocol is registered with PROSPERO [registration number CRD42022320343].^20^

### 2.2 Eligibility criteria

Articles were included in the review if they were written in English, pertained to an adult (18 years of age or over) population, were clinical trials, review articles, studies or point prevalence studies with a publication date between January 2017 and December 2021, related to the AMS strategy IVOS and with specified IVOS criteria.

Studies were excluded if they were case reports and non-studies e.g., conference abstracts, editorials, letters, notes, or related to one particular antimicrobial or specific infection (to exclude non-generalisable criteria).

### 2.3 Information sources

Literature search terms were devised alongside an experienced UKHSA Knowledge and Evidence Specialist (CD). Systematic searches were conducted in OVID Embase and OVID Medline databases. For the initial searches, no start date limit was set, and databases were searched up to 15 December 2021.

### 2.4 Search strategy

The defined research question was: What are the criteria necessary to achieve a safe and effective antimicrobial intravenous-to-oral switch in the hospital inpatient adult population?

See **Table 1** for database search strategies.

**Table 1.**
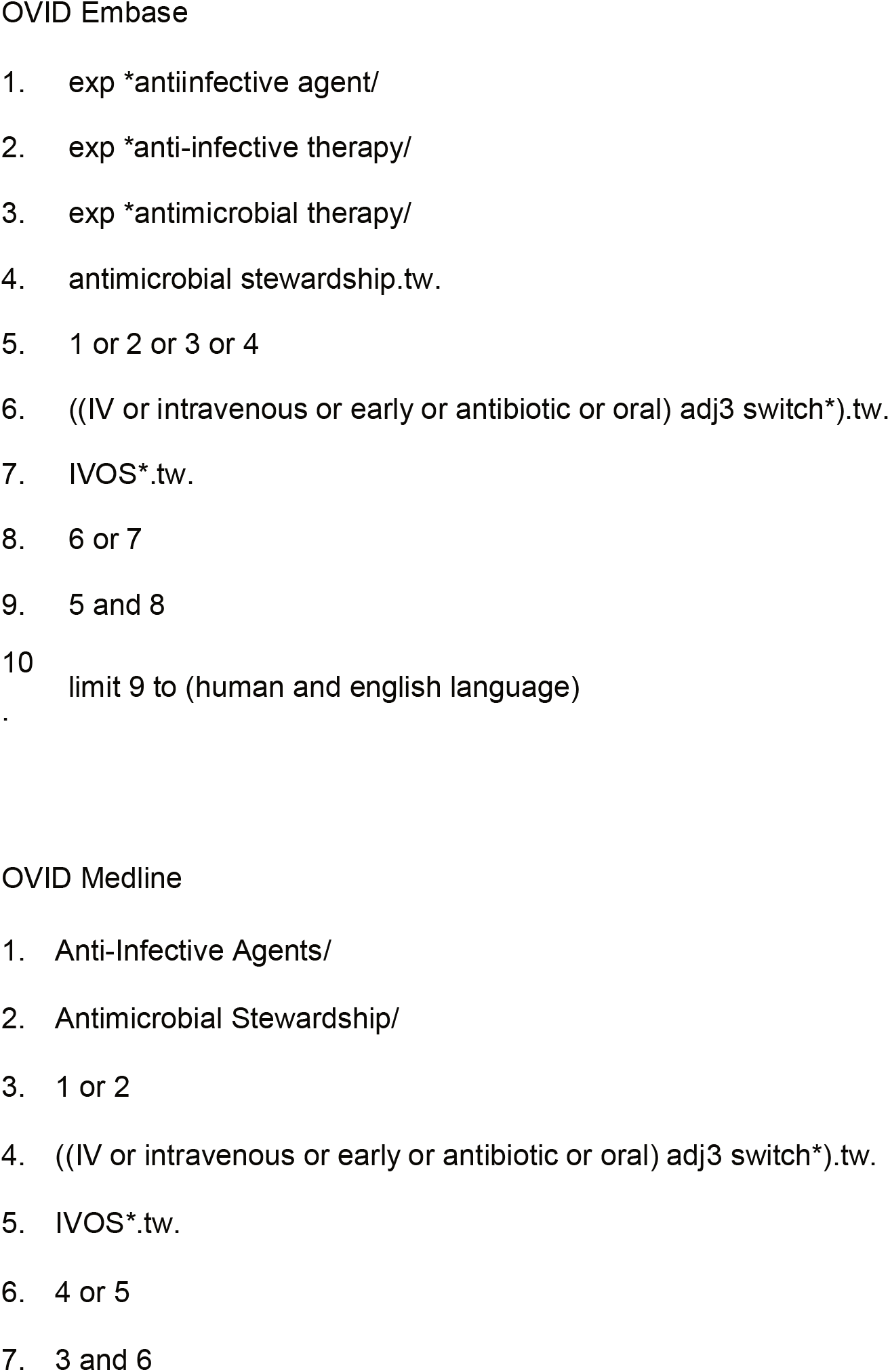
OVID Embase and OVID Medline search strategies

### 2.5 Selection process

Article duplicates were removed on EndNote. One researcher (EH) screened all titles and abstracts for articles relevant to the research question. From this initial screening, Akhloufi *et al*.’s paper published in 2017 was identified as including an IVOS literature review,^21^ thus 2017 onwards became an inclusion criterion to capture evidence published since the most recently identified review. One researcher (EH) screened full articles to ascertain focus of article and identify IVOS criteria. A second researcher (DAO) reviewed the focus themes for accuracy.

A second assessment of a random 20% sample of articles dated between the years 2017 and 2021 was conducted by a third researcher (MM). Discrepancies were resolved between two reviewers (EH, MM) with a third reviewed (DAO) as arbiter.

### 2.6 Data collection process

A data extraction Excel spreadsheet was designed with the following column headings: Authors, Year of publication, Title, Journal, Edition, Pages, Month, Article type, DOI, Keywords, Abstract, URL link, Database, Platform, Language, Continent research conducted in, Country/area, Sample size, Population, Focus, Specifics, Healthcare professional mention, Key findings, Exclude/Include, Reason, Additional comments.

The Focus heading options included AMS (focus on AMS strategies such as IVOS), Antimicrobial (focus on an individual or class of antimicrobial(s)) or Infection (focus on specific infection). Studies with an AMS focus would further be classified under the Specifics column as General (non-IVOS AMS strategy) or IV Switch (IVOS AMS strategy). The studies in the IV Switch column were further evaluated to ascertain if they included IVOS switch criteria.

### 2.7 Data items

The primary outcome sought from the literature was safety of antimicrobial IVOS, defined by measurable IV switch criteria for adults (to include review time, clinical signs and symptoms, infection markers, enteral route, infection exclusions).

Additional outcomes were reduced healthcare associated infections, reduced catheter-related infections, reduced hospital length of stay, reduced costs (e.g., drug equipment), reduced staff workload, reduced IV administration errors, increased patient comfort and mobility, improved AMS practice.

### 2.8 Study risk of bias assessment

The included studies were not assessed for internal validity with a risk of bias tool. This differs to the protocol registered on PROSPERO. This decision was agreed by all co-authors and the reasons are: 1) as a rapid review the risk of bias can be omitted,^22^ and 2) risk of bias tools become more relevant for studies assessing effectiveness of interventions, whereas this review aimed to identify IVOS criteria to subsequently inform a Delphi consensus-gathering process.

### 2.9 Synthesis methods

There was heterogeneity of study designs and outcome measures. This was expected and was not investigated as outside scope of the rapid review. A framework synthesis on included studies was completed.

#### 2.9.1 Development of framework

The framework was derived based on an analysis of IVOS criteria obtained from a sample of UK acute hospital IVOS policies sought though stratified sampling. The target number of IVOS policies was 41 (25%), to include polices from English Trusts and Scottish Health Boards and Northern Ireland and Wales all-country policies. Hospitals from England were selected from an ‘All England Trusts and Trust Types’ UKHSA datasheet by hospital type (acute – small, acute – medium, acute – large, acute – multi-service, acute – specialist, acute – teaching) and region (East of England, London, Midlands, North East and Yorkshire, North West, South East, South West) to ensure representative sampling. English IVOS policies were obtained through internet search or MicroGuide application access. IVOS policies from Scotland’s Health Boards were obtained through internet search, and from Northern Ireland and Wales through internet search and email correspondence. An Excel spreadsheet was designed to collate criteria under headings to inform framework sections.

### 2.10 Certainty of evidence

All included studies were peer-reviewed articles, enhancing the certainty of the evidence. Studies were not formally graded. High evidence rating [H] was assigned to high quality evidence such as clinical trials and reviews, and medium evidence rating [M] was assigned to lower quality studies such as prospective studies and point prevalence studies.

## 3 Results

### 3.1 Framework synthesis

Forty-two acute hospitals (28%, n=148) from England were selected for review of their IVOS policies, two Health Boards (14%, n=14) from Scotland and the All-Wales policy from Wales. No IVOS policy from Northern Ireland was identified at the time of the study period. Overall, 45 (27%, n=164) acute hospital IVOS policies from three UK nations were included for review. IVOS criteria were collated on an Excel spreadsheet under the following 5 framework sections and headings: Review IV within (hours) (Section 1: Timing of IV antimicrobial review); Clinical signs and symptoms (Section 2: Clinical signs and symptoms); Temperature, Heart rate, Blood pressure, Respiratory rate, White cell count, C-reactive protein (Section 3: Infection markers); Gut function, Drug interactions with oral therapy, Allergy to oral therapy (Section 4: Enteral route); and Infection exclusions (Section 5: Infection exclusions) (**Supplementary Table S2**).

### 3.2 Study selection

Four hundred seventy-seven papers were identified from the literature search. Fifty-nine were non-studies (e.g., letters, notes, conference abstracts) and therefore excluded. Akhloufi *et al*.’s IVOS review in 2017^21^ led to the timeframe of included papers to be between 2017 and 2021, excluding a cfurther 308 papers. The remaining 110 articles were screened for relevance to the research question by title and abstract. Seventy papers were excluded due to a focus other than AMS. Forty papers were found to be eligible for inclusion and screened by full article. A further 24 papers were excluded as they contained no IVOS criteria or pertained to the paediatric population (n=1). Sixteen papers with IVOS criteria were included for review. See **Figure 1** for PRISMA flowchart.

**Figure 1.**
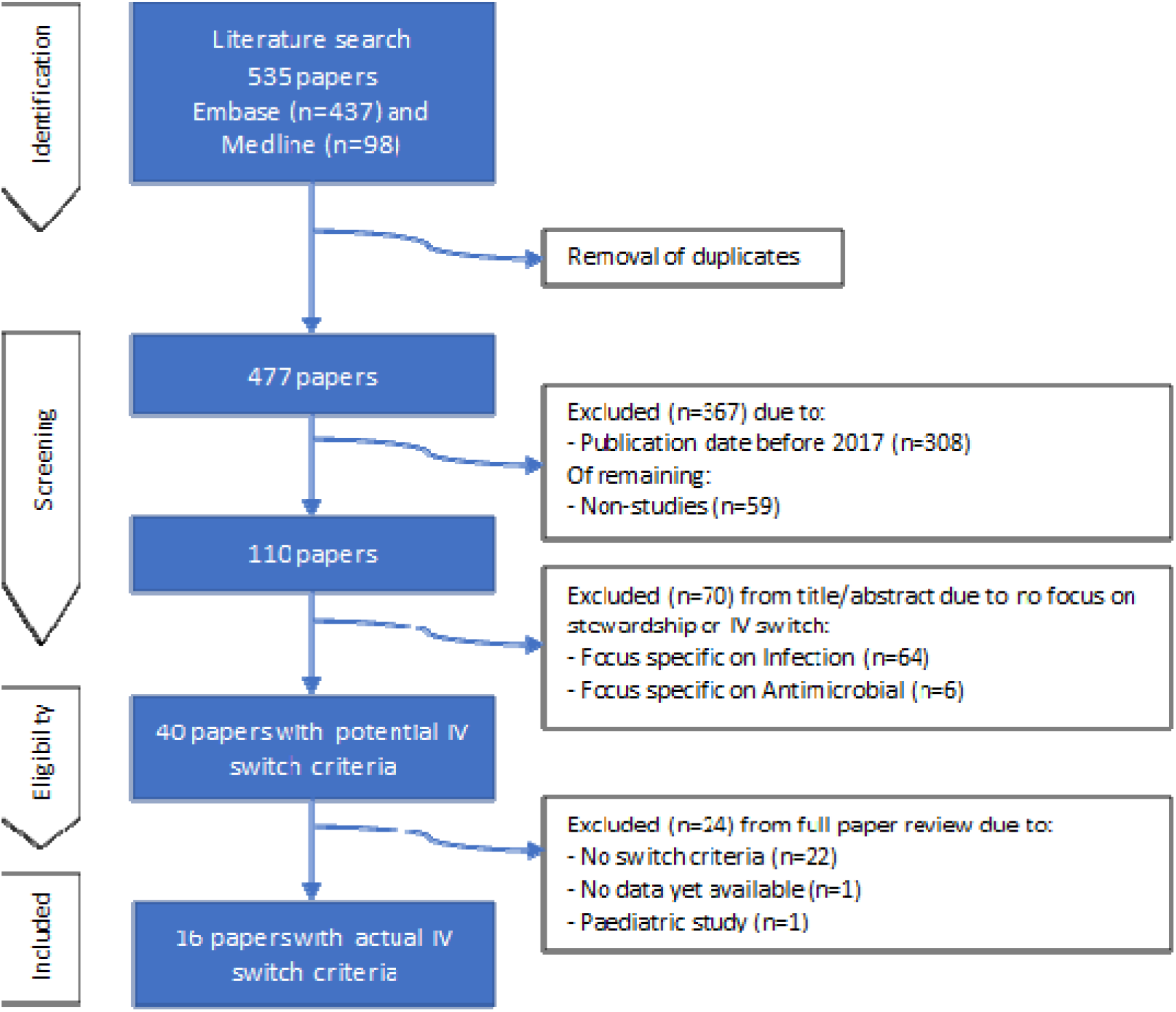
PRISMA flowchart of the literature search and rapid review

The primary outcome of safety of antimicrobial IVOS was identified in eight of the included papers (50%),^12, 21, 23-28^ with remaining papers focusing on outcomes classed as additional for the purposes of this study. All papers included measurable IVOS criteria.

### 3.3 Study characteristics

Most published papers were from 2021 (n=4) and 2018 (n=4). The Netherlands had published the most peer-reviewed articles (n=6), followed by two each from the United Kingdom and Australia, and one each from Canada, India, Malaysia, Switzerland, Thailand and Vietnam. Eight papers presented IVOS criteria for use in clinical practice in table format, two in flowchart format and one as text. The remaining five articles did not state how IVOS criteria were presented. The majority (n=11) had a Switch approach, 4 were unknown and one had a Continue approach (**Table 2**). Switch approach referred to criteria being expressed in terms of ‘if patient meets x criteria, consider switch from intravenous to oral therapy’, instead of the Continue approach of ‘if patient does not meet x criteria, consider continuing intravenous therapy’.

**Table 2.**
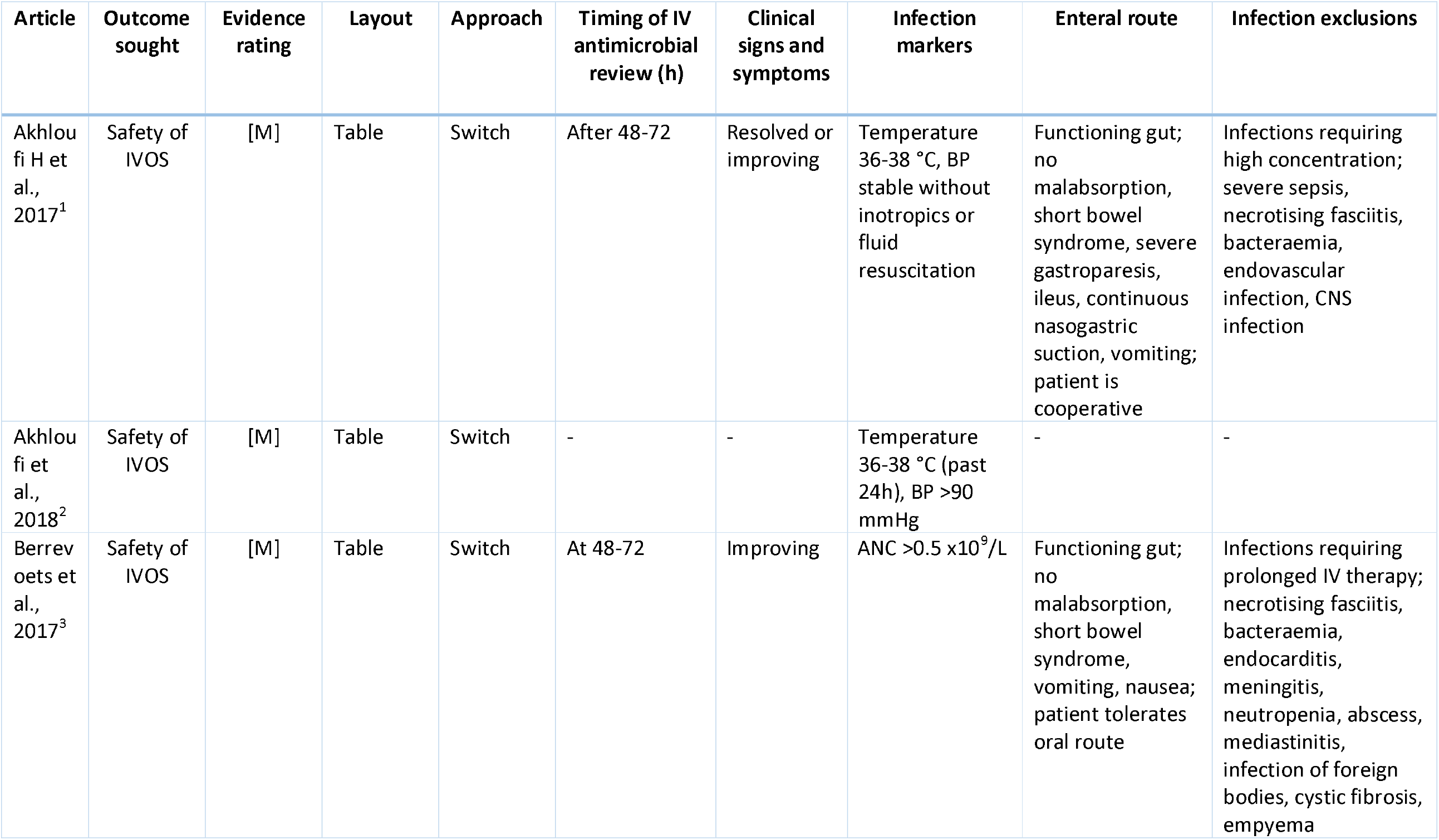
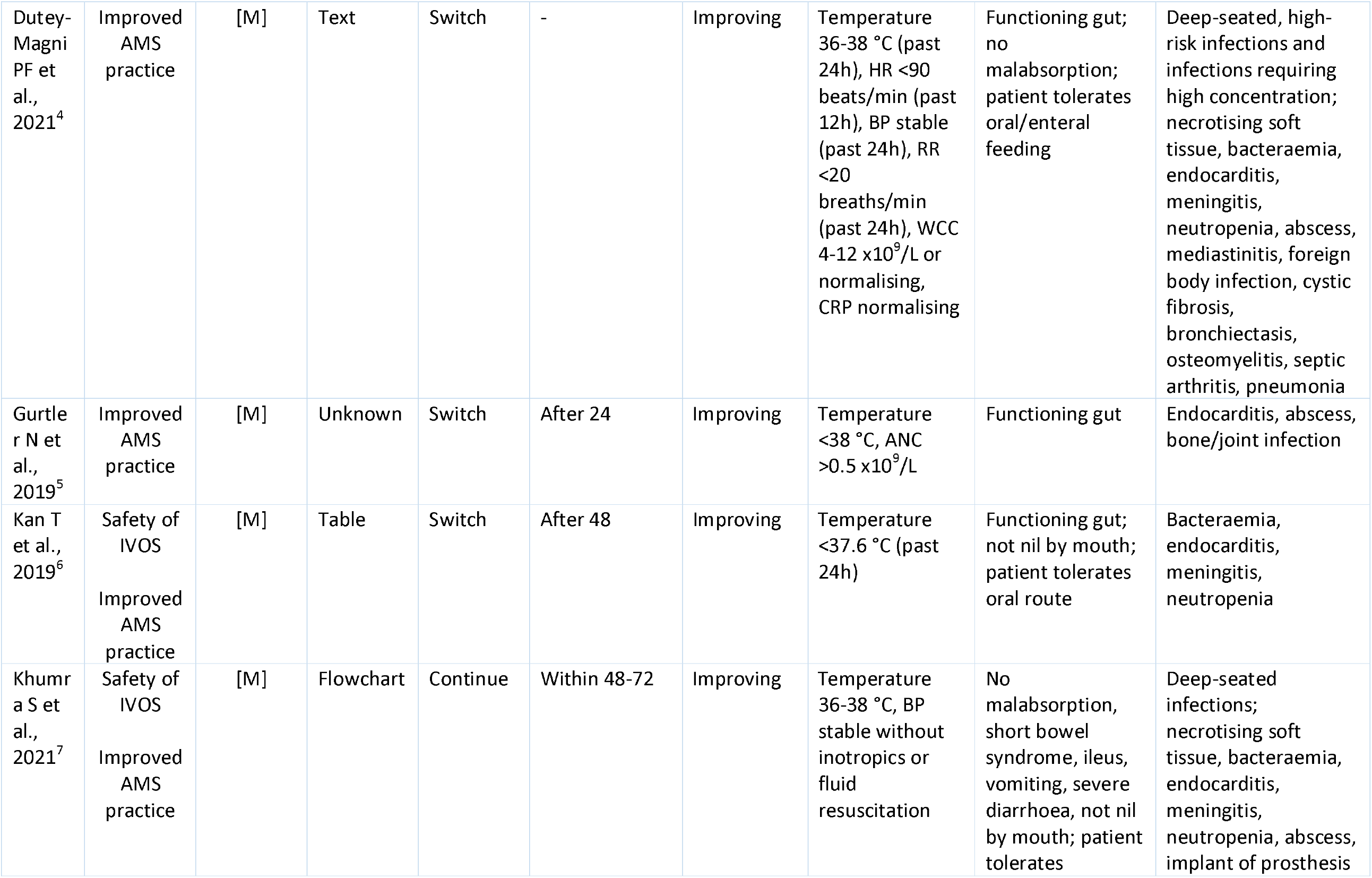
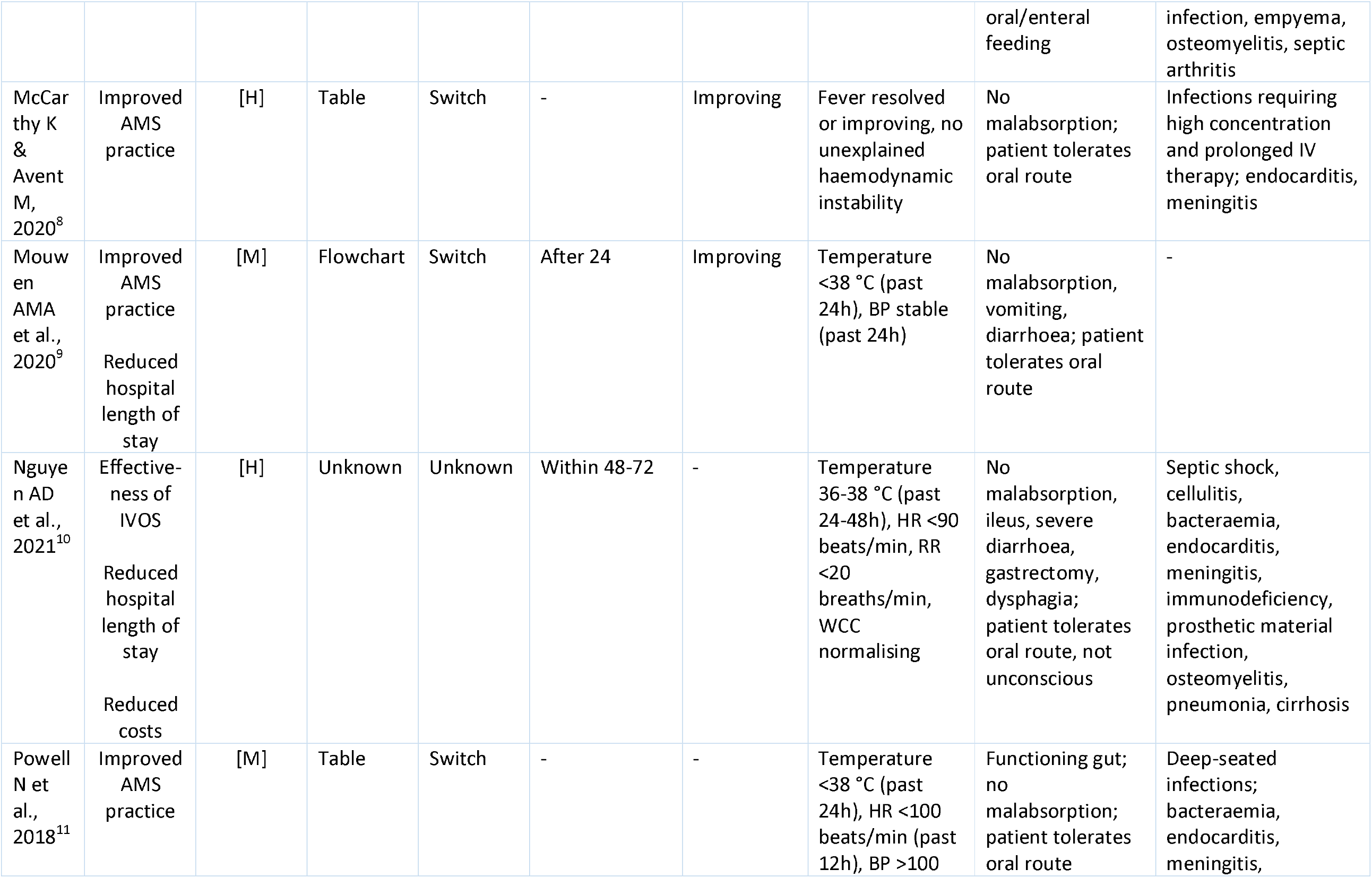
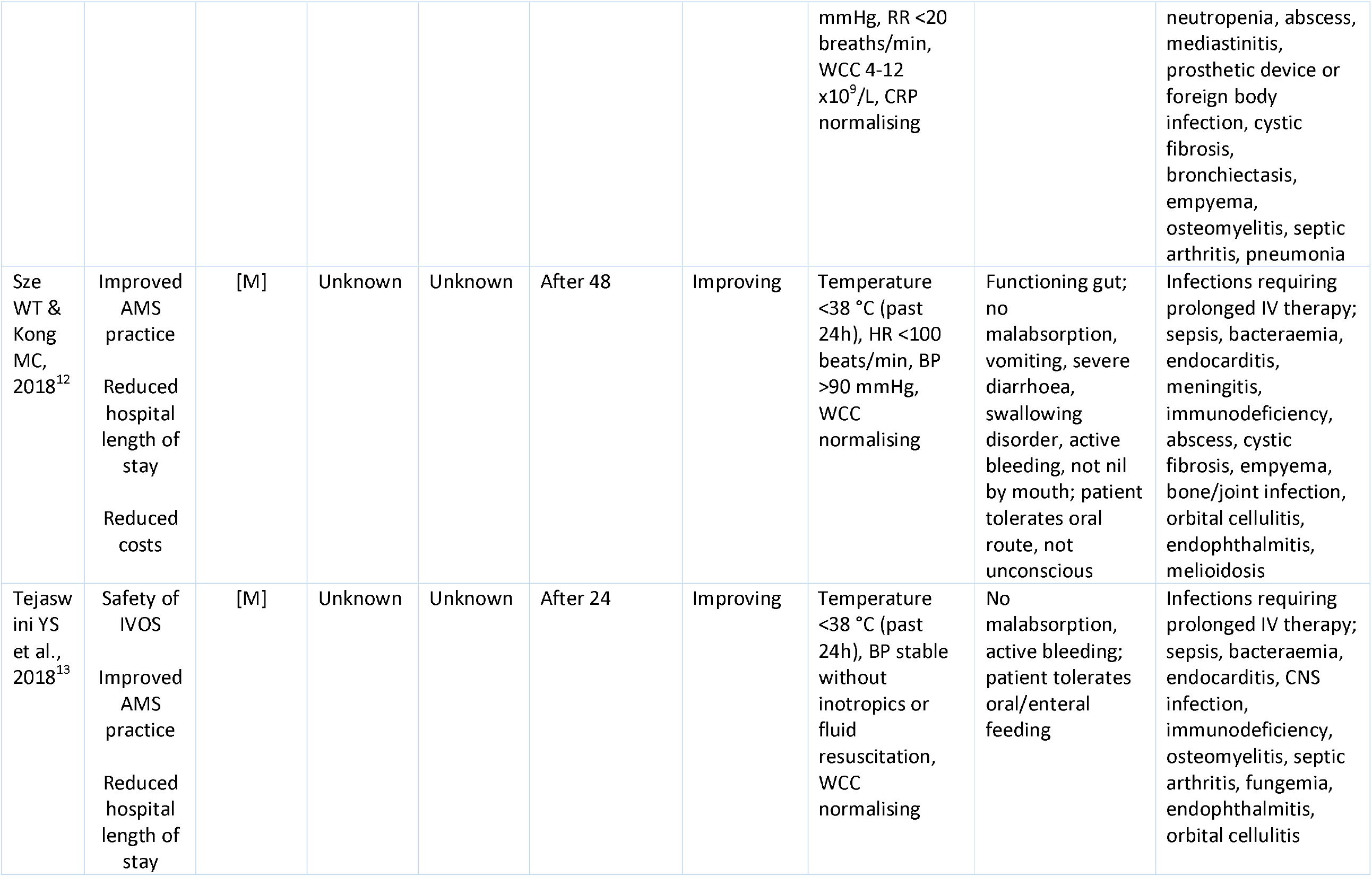
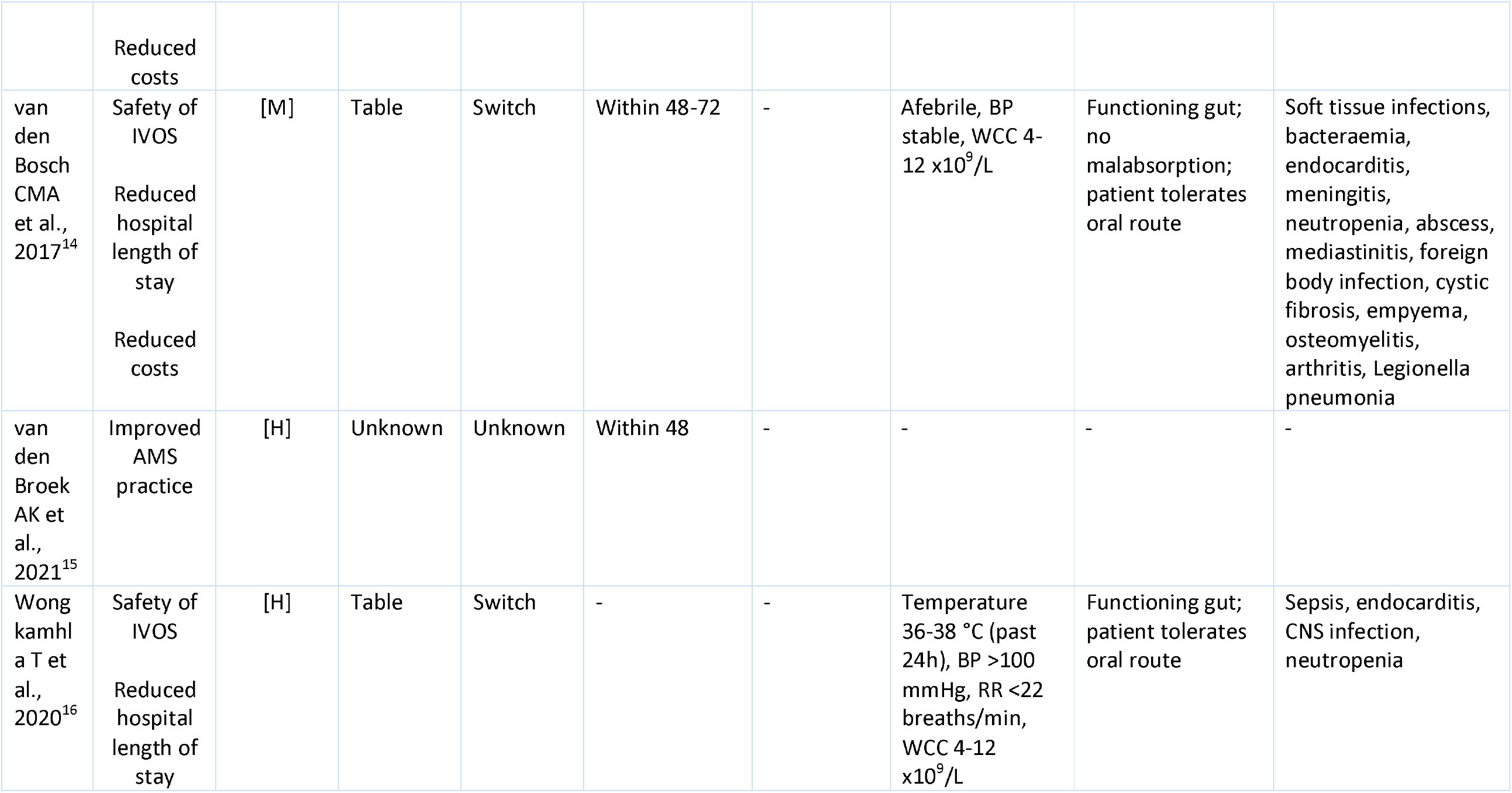
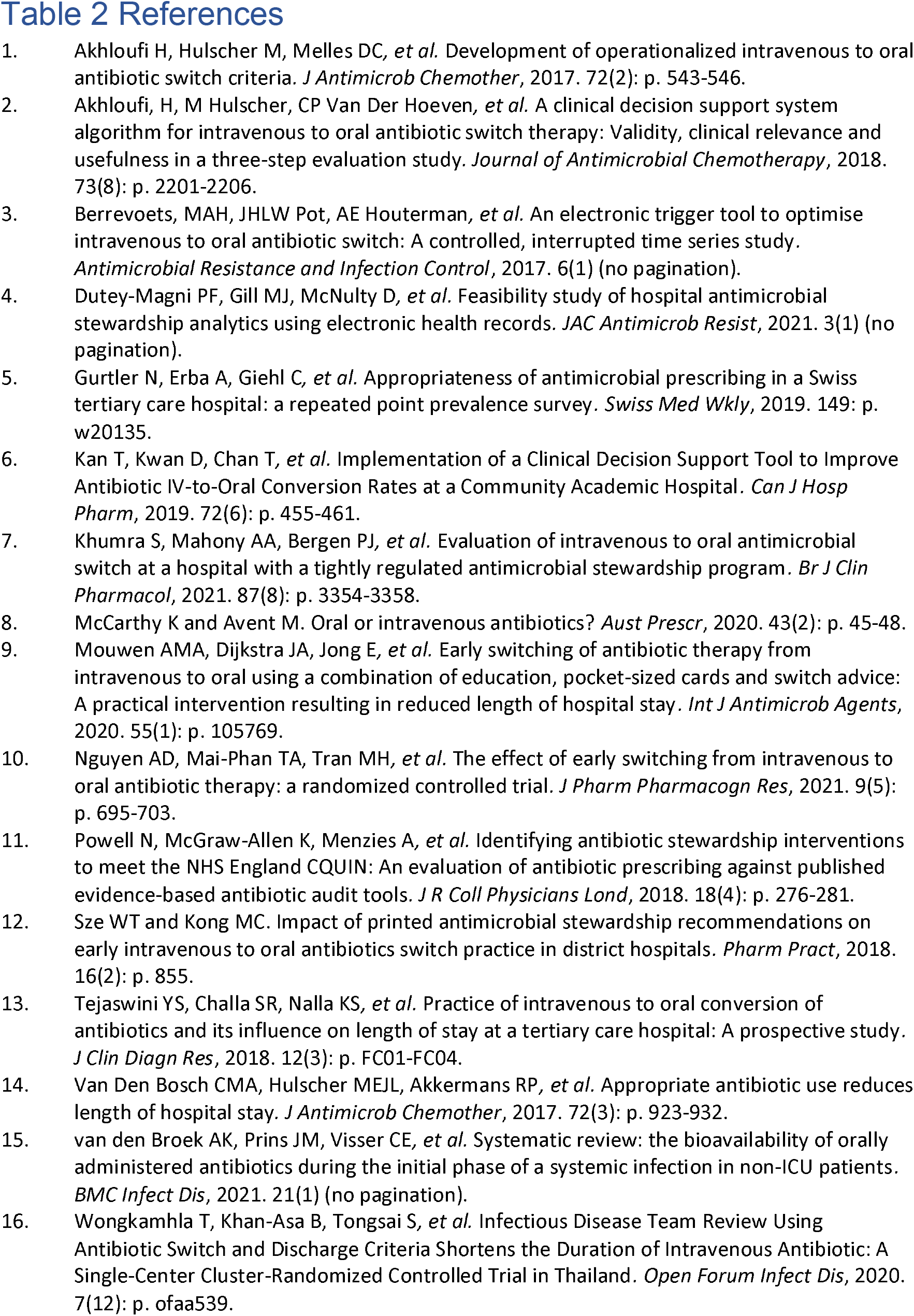
Summary of results of individual studies

### 3.4 Results of individual studies

**Table 2** includes a summary of results of individual studies, including identified IVOS criteria.

### 3.5 Results of syntheses

Eight studies (50%) specified criteria for all 5 sections of the defined framework. Criteria with highest, or over 50%, appearance in the literature, informed by evidence rating was collated. Overall, 33 criteria were identified to go forward into the Delphi process (**Table 3**).

**Table 3.** Criteria from rapid review (n=33) to go forward into Delphi process

| Criteria | Number of papers<br>n, (%) | Evidence rating |
| --- | --- | --- |
| **1. Timing of IV antimicrobial review** |  |  |
| a. Review antimicrobial within **48 hours** | 1 (6) | [H] |
| b. Review antimicrobial within **48-72 hours** | 3 (19) | [M] x2, [H] |
| **2. Clinical signs and symptoms** |  |  |
| a. Clinical signs and symptoms should be improving | 9 (56) | [M] x8, [H] |
| **3. Infection markers** |  |  |
| a. Temperature should be between 36-38°C | 2 (13) | [M] x2 |
| b. Temperature should be between 36-38 °C past **24 hours** | 3 (19) | [M] x2, [H] |
| c. Heart rate should be below 90 beats per minute | 1 (6) | [H] |
| d. Heart rate should be below 90 beats per minute for past **12 hours** | 1 (6) | [M] |
| e. Blood pressure should be stable | 1 (6) | [M] |
| f. Blood pressure stable for past **24 hours** | 2 (13) | [M] x2 |
| g. Respiratory rate should be below 20 breaths per minute | 2 (13) | [M], [H] |
| h. Respiratory rate should be below 20 breaths per minute for past 24 hours | 1 (6) | [M] |
| i. White cell count should be normalising | 3 (19) | [M] x2, [H] |
| j. White cell count should be between 4 and 12 x10<sup>9</sup>/L | 3 (19) | [M] x2, [H] |
| k. White cell count should be between 4 and 12 x10<sup>9</sup>/L or normalising | 1 (6) | [M] |
| l. C-reactive protein should be normalising | 2 (13) | [M] x2 |
| **4. Enteral route** |  |  |
| a. Gastrointestinal tract must be functional | 9 (57) | [M] x8, H |
| b. Patient can tolerate/ swallow oral option | 9 (57) | [M] x6, [H]x3 |
| c. No evidence of malabsorption | 11 (69) | [M] x9, [H] x2 |
| d. No vomiting | 5 (31) | [M] x5 |
| **5. Infection exclusions** |  |  |
| a. Deep-seated infections | 3 (19) | [M] x3 |
| b. Infections requiring high tissue concentration | 3 (19) | [M] x2, [H] |
| c. Infections requiring prolonged IV therapy | 4 (25) | [M] x3, [H] |
| d. Critical infection with high risk of mortality | 1 (6) | [M] |
| e. Endocarditis | 12 (75) | [M] x9, [H] x3 |
| f. Meningitis | 9 (56) | [M] x7, [H] x2 |
| g. Bacteraemia, including *Staph. aureus* | 9 (56) | [M] x9 |
| h. Immunocompromised | 3 (19) |  |
| i. Abscess | 7 (44) | [M] x7 |
| j. Severe or necrotising soft tissue infections | 5 (31) | [M] x5 |
| k. Infections of foreign bodies | 6 (38) | [M] x5, [H] |
| l. Osteomyelitis | 6 (38) | [M] x5, [H] |
| m. Septic arthritis | 5 (31) | [M] x5 |
| n. Empyema | 5 (31) | [M] x5 |

#### 3.5.1 Timing of intravenous antimicrobial review

Eleven of the 16 included studies (69%) contained criteria pertaining to the timing of IV antimicrobial review.

Five of the studies (30%) stated that a review should take place either *within* (n=3), *at* (n=1) or *after* (n=1) the 48–72-hour window from initiation of IV antimicrobial. A clinical trial [H]^9^ and a further two studies [M]^26, 28^ defined early switch as IVOS taking place within the 48-72 hours.

Three studies (19%) stated that a review must occur *within* (n=1) or *after* (n=2) 48 hours from initiation of IV antimicrobial. A systematic review [H] investigated early IVOS as being within 48 hours.^29^ Three studies (19%) [M] recommended IVOS review to occur after 24 hours.^27, 30, 31^

#### 3.5.2 Clinical signs and symptoms

Ten of the 16 studies (63%) included clinical signs and symptoms as a criterion for early IVOS. Nine studies (56%) stated that clinical signs and symptoms must be improving; one of these studies was a (non-systematic) review article [H]. A variance in criterion wording was captured by one study that stated that clinical signs and symptoms must be resolved or improving for safe IVOS.^21^

#### 3.5.3 Infection markers

The infection markers identified in the literature were temperature,^9, 12, 21, 23, 25-28, 30-35^ heart rate,^9, 32-34^ blood pressure,^12, 21, 23, 26-28, 31-35^ respiratory rate,^9, 12, 32, 33^ white cell count ^9, 12, 24, 27, 28, 30, 32-34^ or absolute neutrophil count^24, 30^ and C-reactive protein.^32, 33^

##### 3.5.3.1 Temperature

Temperature was the most frequently mentioned infection marker, appearing in 14 of the 16 studies (88%). The most common criterion variance was that temperature must be between the 36-38°C range, appearing in 6 studies (38%). Of these 6 studies, three stated that 36-38°C must have been sustained for the past 24 hours – one of these studies was a clinical trial [H],^12^ and one stated for the past 24-48 hours, also a clinical trial [H].^9^ Six studies (38%) [M] did not state a temperature range, but instead suggested that it must be less than 38°C,^27, 30, 31, 33, 34^ or less than 37.6°C.^25^ Two studies (13%) did not specify a temperature at all, but instead commented on the need for patient’s fever to be resolved or improving,^35^ or patient to be afebrile.^28^

##### 3.5.3.2 Heart rate

Four of the 16 studies (25%) included heart rate (HR) as an infection marker for safe IVOS. Two of them (12.5%) stated HR must be below 90 beats/minute – the clinical trial [H] provided no timeframe,^9^ whilst the study [M] stated HR must have been sustained for the past 12 hours.^32^

##### 3.5.3.3 Blood pressure

Eleven of the 16 studies (69%) included blood pressure (BP) as an infection marker for early IVOS. The most common wording for the criterion was BP needed to be stable (n=3, 19%),^28, 31, 32^ and BP needed to be stable without inotropics or fluid resuscitation (n=3, 19%).^21, 26, 27^

##### 3.5.3.4 Respiratory rate

Four of the 16 studies (25%) included respiratory rate (RR) as an infection marker for IVOS. Three studies (19%) stated RR must be below 20 breaths/minute; two of them had no associated timeframe,^9, 33^ of which one was a clinical trial [H],^9^ and the third, a study [M], stated that the specified RR must have been sustained for the past 24 hours.^32^

##### 3.5.3.5 White cell count

Nine of the 16 studies (56%) included white cell count (WCC) or absolute neutrophil count within criteria for early IVOS. The preferred wording was divided between WCC must be normalising (n=3, 19%)^9, 27, 34^ and WCC must be within 4-12 x10^9^/L (n=3, 19%).^12, 28, 33^ Each of these criterion wordings appeared in clinical trials [H].^9, 12^

##### 3.5.3.6 C-reactive protein

C-reactive protein (CRP) was the least included infection marker, appearing in only 2 of the 16 studies (13%). Both studies [M] stated that CRP must be normalising for a safe IVOS.^32, 33^

#### 3.5.4 Enteral route

Fourteen of the 16 included studies (88%) had criteria that related to the enteral route. The most common criteria in the literature was that there must be no malabsorption, appearing in 11 of the 16 studies (69%), of which one was a clinical trial [H]^9^ and one a review [H].^35^

Nine studies (56%) stated that the gut must be functioning and 5 studies (31%) stated the patient must not be vomiting.

#### 3.5.5 Infection exclusions

This section included both general information (e.g., infections requiring prolonged IV therapy, deep-seated infections) as well as specifically mentioned infections. The infections with highest counts in the literature were endocarditis (n=12, 75%) including in two clinical trials [H]^9, 12^ and one review^35^ and meningitis (n=8, 50%). Infections with lowest appearance in the literature included mediastinitis (n=4, 25%) and bone and joint infections (n=2, 12.5%).

## 4 Discussion

Early IVOS has numerous advantages compared to continuing intravenous therapy in patients that may be eligible for oral administration. The risk of cannula-related infections and thrombophlebitis would reduce, as would the costs of equipment and staff time. Patient comfort and mobility would be promoted, and the absence of intravenous antimicrobial infusions or injections could result in earlier patient discharge from hospital.^36^ Analysis of individualised hospital IVOS policies showcased the variety of IVOS criteria in clinical use. The standardisation of healthcare practice benefits patient safety and outcomes, however it must be evidence-based.^37^ Through this rapid review, the literature was systematically searched and 33 IVOS criteria were identified as part of a 5-section framework.

### 4.1 Criteria sections

#### 4.1.1 Timing of intravenous antimicrobial review

The most common timing stated in the literature for IVOS review from initiation of antimicrobial intravenous therapy was 48-72 hours, specifically within the 48–72-hour window. Prior guidance outlines the need to review patients’ clinical diagnosis and ongoing need for antibiotics at 48-72 hours.^38, 39^ In the UK national Start Smart – Then Focus AMS Toolkit,^38^ switching antibiotics from intravenous to oral is one of five recommended choices, alongside stopping antibiotics, changing to a narrower or broader spectrum antibiotic, continuing antibiotic treatment stating time of next review or considering Outpatient Parenteral Antibiotic Therapy. Studies have since questioned whether an earlier review, such as review within 48 hours of intravenous initiation, is warranted.^29, 40^ Van den Broek *et al*. commented that theoretical reasons for an early switch (even within 24 hours) exist; for example, if patient has an intact gastrointestinal tract and the oral antibiotic option has adequate bioavailability (generally data obtained from studies carried out in healthy and/or critically ill patients). However, improved knowledge on antimicrobial bioavailability in the acute stage of infectious illness, when the patient may be febrile and drug pharmacokinetics more unfamiliar, would further strengthen the case for earlier IVOS.^29^

#### 4.1.2 Clinical signs and symptoms

Signs as objective manifestations of infectious disease, and symptoms as subjective manifestations,^41^ underline the importance of noting improvement, or deterioration, in both the observed response to infection and its management as well as the self-reported accounts of the patient. This rapid review showcases evidence to suggest that the improvement of a patient’s signs and symptoms is an important criterion to include for safe IVOS.^21, 24-27^

#### 4.1.3 Infection markers

Temperature was the predominant infection marker criterion across all studies, with higher evidence for stating a temperature range compared to stating that temperature must be below a certain figure. Infection can elicit thermoregulation in the host, increasing body temperature (fever/hyperthermia) or decreasing it (hypothermia) as in the case of sepsis,^42^ thus corroborating the importance of including a temperature range in IVOS criteria.

Despite temperature being a vital marker of infection, normothermia, categorised as 36.1-38°C, has been associated with infection and higher patient mortality than hyperthermia.^43^ Normothermia therefore does not exclude the need for antibiotics; and if antibiotics are given, normothermia would not necessarily mean that a switch from intravenous to oral therapy should be made. Other infection markers, to include physiological and inflammatory parameters,^43^ must be considered to ensure any IVOS decision is safe.

The National Early Warning Score (NEWS) developed by the UK Royal College of Physicians is a recognised clinical assessment tool for use in acute hospital and ambulance settings. It was first published in 2012 with a second version, NEWS2, released in 2017. The tool ‘improves the detection and response to clinical deterioration in adult patients and is a key element of patient safety and improving patient outcomes’.^13, 44^ NEWS2 aggregates individual scores from the following physiological parameters: respiration rate, oxygen saturation, systolic blood pressure, pulse rate, level of consciousness or new confusion, and temperature. Certain parameters overlap with the IVOS criteria found in the literature, notably temperature, but also respiration/respiratory rate and blood pressure. The marker named heart rate in this rapid review is specified as pulse rate in NEWS2.

For each marker of HR, BP and RR, the literature showcased a variety of nuances in criteria wording. Some criteria, but not all, specify marker values with or without a particular timeframe for which the specified infection marker must be sustained for; e.g., HR below 90 beats/min for the past 12 hours. The benefit of the NEWS2 score is that ranges, not merely single values, are used to assess clinical deterioration. Additionally, it is a tool endorsed for use in patients with acute infection or at risk of infection and widely implemented across UK NHS hospitals.^13^ Presenting separate criteria for HR, BP and RR is one way to achieve safe IVOS,^12, 21, 23, 26-28^ however presenting them combined into a NEWS2 scoring criterion would offer an alternative evidence-based way to achieve safe IVOS. Other countries have also adopted Early Warning Scores (EWS) to identify patients at risk of deterioration, especially from infection, thus IVOS criteria in other countries could consider EWS as a criterion. In Canada, for example, the Hamilton Early Warning Score of similar prognostic accuracy to NEWS2 is used.^45^ In the United States of America, a variety of tools exist with the NEWS tool considered most accurate to identify patients at high risk of mortality.^46^

#### 4.1.4 Enteral route

This section of the framework is concerned with maximising enteral antibiotic absorption from the gastrointestinal tract (GIT), and hence maximising antibiotic bioavailability and therapeutic action. Antibiotic administration via the intravenous route achieves 100% bioavailability.^36^ Some antibiotics are known to have good oral bioavailability, for example clindamycin, doxycycline, linezolid and metronidazole have over 90% oral bioavailability.^36, 47^

The literature outlines numerous criteria to ensure a switch to oral administration, or indeed to enteral administration to include use of enteral tubes, achieves as close to 100% bioavailability as practicable. The criteria centre on patient characteristics, predominantly in relation to their GIT function e.g., no malabsorption present, but also in relation to their level of alertness e.g., patient is not unconscious. The latter resonates with one of the physiological scores included in the NEWS2 (level of consciousness or new confusion scoring),^13^ further promoting inclusion of EWS as a combination criterion for safe IVOS.

The literature includes no mention of antibiotic pharmacological properties such as antibiotic bioavailability, nor the need to review drug interactions between oral antibiotic options and any other medication the patient may be taking or to check for allergies. It is worth noting that these criteria do appear in the sample of acute hospital IVOS policies used to inform the framework analysis (**Supplementary Table S2**).

#### 4.1.5 Infection exclusions

Over the years, randomised controlled trials have been undertaken comparing treatment arms of prolonged intravenous antibiotic courses and shorter courses. The research has demonstrated equal outcomes between treatment arms for infections such as community-acquired pneumonia and neutropenic fever.^48^ These results have enabled change in clinical practice and advancement in AMS.

However, not all infections have the evidence to make them eligible for early IVOS. This section of the framework provides a caveat around certain infections when making early IVOS decisions. Endocarditis was the infection exclusion with highest appearance in the literature. The POET trial enrolled patients with left-sided endocarditis and compared full course intravenous therapy versus oral switch after a minimum of 10 days intravenous therapy. IVOS was found to be noninferior in terms of treatment failure at one year compared to full intravenous therapy.^49^ However, the timing of IVOS (after at least 10 days of intravenous therapy) was much later than the timing for IVOS proposed in the included literature for this rapid review. Considering the current available evidence, endocarditis may remain as an infection exclusion within IVOS criteria.

Bone and joint infections appeared only twice in the literature, in papers published in 2018^34^ and 2019.^30^ Around that time, the OVIVA trial investigated early IVOS for bone and joint infections and concluded that oral antibiotic treatment was noninferior to intravenous treatment. Trial IVOS decisions were led by specialist teams,^50^ so it remains to be said that in the standardisation process of IVOS criteria, which also serves to equip non-specialist teams, professional judgment regarding when to refer patients for specialist input remains important in IVOS decision-making.

### 4.2 Strengths and limitations

A strength of this study includes a systematic literature search being undertaken to capture papers relevant to IVOS AMS strategy. Papers were not limited by country or region, thus the criteria identified can serve as a starting point for any institution globally. Two researchers (EH, DAO) were involved in reviewing themes and included papers, with a separate researcher (MM) undertaking a second assessment of a random sample of papers.

A limitation is that non-English papers were excluded, potentially discounting research on IVOS criteria conducted in non-English speaking nations. During the study period, the authors were not aware of a published validated rapid review reporting protocol.^51^ However, the WHO and Cochrane approaches to rapid reviews were taken into consideration and the authors followed through the PRISMA reporting guidelines.

## 5 Conclusion

The benefits of early IVOS include reduced risk of healthcare-associated and catheter-related infections, reduced costs, staff workload and hospital length of stay, and increased patient mobility and comfort. Individual hospital polices contain variable IVOS criteria with an unknown evidence-base. This rapid review has identified and collated 33 evidence-based IVOS criteria from the literature and presented them within 5 distinct and comprehensive sections.

Further research is required to achieve a national consensus on IVOS criteria from healthcare professionals providing the care for hospitalised adult patients and making the decisions regarding infection management. In the acute hospital setting, operationalisation of IVOS criteria as a tool to promote best practice needs to be explored and must take into consideration the roles and strengths that a multidisciplinary team brings to AMS.

## Data Availability

All data relevant to the study are included in the article or uploaded as supplementary information.

## 6 Author contributions

Conceptualisation: DAO, EH. Methodology: EH undertook framework analysis, literature review and drafted paper, CD contributed to literature searches, MM undertook the random second assessment of articles, DAO validated aspects of methodology and supervised the review. All authors reviewed and approved final version.

## 8 Declarations

None to declare.

## 9 Ethics approval

This study does not involve human participants.

## 10 Data availability statement

All data relevant to the study are included in the article or uploaded as supplemental information.

